# Ethical Issues in Residency Education Related to the COVID-19 Pandemic: A Narrative Inquiry Study

**DOI:** 10.1101/2023.01.27.23285063

**Authors:** Aliya Kassam, Stacey Page, Julie Lauzon, Rebecca Hay, Marian Coret, Ian Mitchell

## Abstract

**Background:** Amidst the pandemic, residency programs were faced with new challenges to provide care and educate junior doctors (resident physicians). We sought to understand both the positive and negative experiences of first-year residents during COVID-19, as well as to describe potential ethical issues from their stories.

**Method:** We used narrative inquiry (NI) methodology and applied a semi-structured interview guide that included questions pertaining to ethical principles as well as both positive and negative aspects of the pandemic. Sampling was purposive. Interviews were audio-recorded and transcribed. Three members of the research team coded transcripts in duplicate to elicit themes. A composite story with threads was constructed. Discrepancies were resolved through discussion to attain consensus.

**Results:** Eleven residents participated from Internal Medicine (n=2), Family Medicine (n=2), Ophthalmology (n=1), General Surgery (n=1), Pediatrics (n=1), Diagnostic Radiology (n=1), Public Health (n=1), Psychiatry (n=1), Emergency Medicine (n=1). Resident stories had three common themes in which ethical issues were described: 1) *Intersecting healthcare and medical education systems*, 2) *Public health and the public good*, 3) *Health systems planning/healthcare delivery*.

**Discussion:** The pandemic exacerbated the lack of autonomy experienced by resident physicians. The notion of public health and the public good at times eclipsed individual wellbeing for residents and patients alike.

**Conclusion:** Efforts to understand how resident physicians can be engaged in their own education as well as how they can navigate public health crises with respect to ethical principles could benefit both residency education and healthcare delivery.

## Introduction

Ethical issues that arise during pandemic health emergencies understandably impact healthcare providers and others who have responsibilities to the public. The COVID-19 pandemic brought along with it complex ethical issues and it is important to recognize there is a tension between the personal limits of healthcare providers and the maintenance of public service responsibilities. This tension coupled with the associated ethical issues present significant implications for healthcare provider wellbeing. ^1 2^

In 1984, Andrew Jameton defined moral distress as: a situation “when one knows the right thing to do, but institutional constraints make it nearly impossible to pursue.” ^3^ While healthcare providers encounter ethical issues on a daily basis regardless of the pandemic, within the context of COVID-19 different constraints have included conflicting policies, deviations from usual care, and decisions based on scarce resource allocation. ^4^ Residency training programs, which are often integral parts of healthcare delivery have experienced changes in content, format, and process. ^5 6^

Prior to the pandemic, residents were expected to be in-person to maximize training opportunities for assessment of competence. Additionally, clerks (senior medical students) also took part in in-person training with patients (clerkship) to ensure maximal training and exposure to multiple specialties. As part of the matching process, this in-person training aided the residents’ decisions when choosing residency programs.

During the beginning of the pandemic, residency programs were faced with new challenges: provide care, educate residents while ensuring learner and patient safety, and allocate resources amidst the suspension of standard in-person learning opportunities. ^5 7^ Resident physicians were in a unique position given that they straddle the worlds of learning within the postgraduate medical education (PGME) system while working within the healthcare system. ^8^ Although this duality in roles existed regardless of the pandemic, the COVID-19 pandemic may have introduced new ethical issues or exacerbated existing ethical issues. ^1 2^

Around the world, the recent shift to competency-based medical education (CBME) from the traditional time-based model is meant to allow for feedback and assessment as residents meet Entrustable Professional Activities (EPAs) ^9^ and milestones. ^10 11^ Given that the COVID-19 pandemic however led to a decrease in resident exposure and experiences within their respective specialities, the concern that residents may not be adequately achieving the EPAs ^9^ set forth by the Royal College of Physicians and Surgeons in Canada (RCPSC) still exists.

The globally recognized Canadian Medical Education Directives for Specialists (CanMEDS) framework created by the RCPSC includes seven roles that address the overarching goal of improving patient care. ^12^ In particular ^12^, the Professional CanMEDS role encompasses ethics and states:

> “The Professional Role reflects contemporary society’s expectations of physicians, which include clinical competence, a commitment to ongoing professional development, promotion of the public good, adherence to ethical standards, and values such as integrity, honesty, altruism, humility, respect for diversity, and transparency with respect to potential conflicts of interest. It is also recognized that, to provide optimal patient care, physicians must take responsibility for their own health and wellbeing and that of their colleagues.”

Ethics education in medical school and residency has focused on biomedical ethics. ^1 13 14^ More specifically, biomedical or clinical ethics conventionally refers to ethics situated within a specific context such as medical education or clinical practice. Biomedical ethics also draws on traditional ethical principles such as autonomy, non-maleficence, beneficence and justice and these principles are applied “during unaccustomed or rare clinical circumstances.” ^15^

Despite effective professionalism curricula in medical education, ethics teaching may not always lead to enhanced ethical reasoning. ^16^ Gaps in teaching any ethics existed prior to the pandemic (11, 12), let alone ethics during a pandemic. ^1 2^ As such, “residents may have little direction on the practical management of ethical issues and principles.” (10) Furthermore, residents “have little experience applying them in complex patient care situations, and they are at a loss when they confront a situation in which at least one important principle of medical ethics must be relinquished.” (10) While the rationale for competency-based medical education is “to ensure physicians graduate with the competencies required to meet local health needs,” ^11^ research has also shown a lack of awareness regarding the commitment to the public aspect of the Professional Role even before the onset of the COVID-19 pandemic. ^17^

Although these gaps in ethics education, a lack of experience with managing ethical issues, and the potential lack of awareness amongst residents regarding broader public health needs all existed prior to the pandemic, the impact of the pandemic on residency training and the adoption of methods to maintain consistency in training quality has added more complexity. ^5^ For example, in the healthcare system changes to usual operations and the allocation of resources such as resident service significantly altered routine workflow in hospitals. ^5^ Amidst these disruptions to training and healthcare, the delicate balance of personal and public safety may have been exacerbated by the pandemic that led to potential ethical issues for resident physicians who may not have had the conceptual tools to deal with them. (15, 16)

Specifically, in public health ethics the entity of concern is not the individual but rather populations^1^, and thus can be considered a form of macroethics thereby preceding the individual patient-physician interaction (microethics). ^18^ Public health by its nature is all about the public good. Second, the prevention of disease and healthcare services must be patient centred. Third, accomplishing public health most often requires government enforcement. ^19 20^ Last, public health entails seeking to avoid poor health outcomes while promoting good health outcomes which can be considered a consequentialist and outcome-oriented foundation of public health. ^19^

Microethics on the other hand, refers to an innovative concept of perpetual inquiry that takes place as part of relational interactions between people daily. ^13 15 18^ An example of this is the patient-physician relationship in the healthcare setting. Microethics refers to ethical issues of common importance which help to promote the quality of healthcare and encompasses relational ethics. ^18^ Microethics, however is still of significance with respect to public health ethics, since it draws on traditional ethical principles that are still relevant to public health. ^13^

Prior to the pandemic, residents in acute care settings may have been working in microethical situations ^1 2^ that may have emphasized commitment to the profession, commitment to patients, and commitment to their own wellbeing as per the Professional Role. There may have been little exposure to commitment to the public (macroethical situations) until the pandemic. ^17^

The circumstances of COVID-19 in residency education described above, such as the suspension of in-person learning as well as re-deployment of resident physicians, may have led to complex ethical issues that called professional ethics and values into conflict for all healthcare providers alike. As Upshur states, “the overarching concern for the individual patient found in clinical ethics is not neatly analogous to a concern for the health of a population. As well, there is no clear analogy to the fiduciary role played by physicians. Simply put, populations are constituted by diverse communities of heterogeneous beliefs and practices. These may at times come into conflict.” ^21^

With multiple factors at play in residency education and service during the COVID-19 pandemic, frameworks and operating standards in both healthcare and residency education evolved quickly. In some parts of the world, resident re-deployment was completely voluntary, and residents were not provided a stipend for their COVID-19 service. Conversely, in other parts of the world, re-deployment was compulsory and a stipend was provided through the government. ^22^

To better understand the impact of the COVID-19 pandemic on junior residents, we sought to explore their experiences during the first wave of the COVID-19 pandemic through their own stories. We also sought to construct ethical issues that may have arisen based on their stories.

## Methods

This qualitative research study used narrative inquiry (NI) to conduct in-depth semi-structured interviews with resident physicians during the first wave of the COVID-19 pandemic. Narrative inquiry can be situated within a constructivist stance alongside interpretivism. ^23^ Narrative inquiry is defined as the study of experiences through an understanding of people’s stories. ^24^ It is a way of conceptualizing and examining experiences. Narrative inquirers encourage story telling as a means to describe experience while following an iterative, reflexive method. ^25^ The researchers sought to use these stories to identify and construct ethical issues that may have occurred during the pandemic’s first wave. Narrative inquiry is based upon experience and the ^26^ “…regulative ideal for inquiry is not to generate an exclusively faithful representation of a reality independent of the knower…. The regulative ideal for inquiry is to generate a new relation between a human being and their environment.” ^27^

Clandinin and Connelly ^24^ state there are three aspects that need to be made explicit with narrative inquiry: *personal* and *social* (Interaction); *past, present, future* (Continuity); and *place* (Situation). More specifically they maintain:

> “Using this set of terms, any particular inquiry is defined by this three-dimensional space: studies have temporal dimensions and address temporal matters: they focus on the personal and the social in a balance appropriate to the inquiry: and they occur in specific places or sequences of places.” ^28^

In this study, personal and social interaction took place by each of the resident participants with the researcher and considered the interaction of residency education (learning and working) before the COVID-19 pandemic, during the COVID-19 pandemic, and the potential impact of COVID-19 more broadly in the future. This was based on the setting or context of the latter part of the first wave of the COVID-19 pandemic in June 2020 in a social distanced manner (space and place).

### Issue of Reflexivity

Throughout the study, first author was aware of potential biases that could have impacted data collection and analysis and was mindful to ensure that the analysis was impartial.

Furthermore, the relationship between, and influence of, the researcher and participant were made explicit in each interview. Given that the data are largely a result of the interaction between the researcher and the participant in interviews, the researcher did not require continual awareness of this relationship. The researcher consistently used memoing to record reflective notes that included what they learned from the data as well as their thoughts on potential biases. ^29^ Furthermore, once the researchers reached consensus on the presentation of the results, the results were sent to the resident participants for member checking.

### Participant Recruitment

Purposive, convenience sampling was used to recruit resident physicians. Resident physicians were recruited by way of two resident physician co-investigators, and through the Office of Postgraduate Medical Education for this study via email and the snowballing technique. Sampling was purposeful to ensure a diverse spread of residency program specialties. The inclusion criteria for the participants were that they were registered in a residency training program and were in their first year of their program to satisfy the role of being considered a junior resident.

### Data Collection

All the authors co-created and reviewed the interview guide which was developed to reflect the microethical and macroethical constructs described above, as well as the four ethical principles of autonomy, beneficence, maleficence, and justice. The fear associated with disclosure of issues that could make learners appear vulnerable has been well established in the literature. ^30–32^ As a result, questions were intentionally kept open-ended and semi-structured to elicit participant stories and avoid censored responses. During the consent process resident physicians were told the study was about ethical issues during the COVID-19 pandemic.

The interview guide is shown in Table 1 below. Interested residents contacted the first author to schedule an interview. Individual telephone interviews were held between May 2020 and July 2020. Interviews were digitally audio recorded and transcribed verbatim.

**Table 1.**
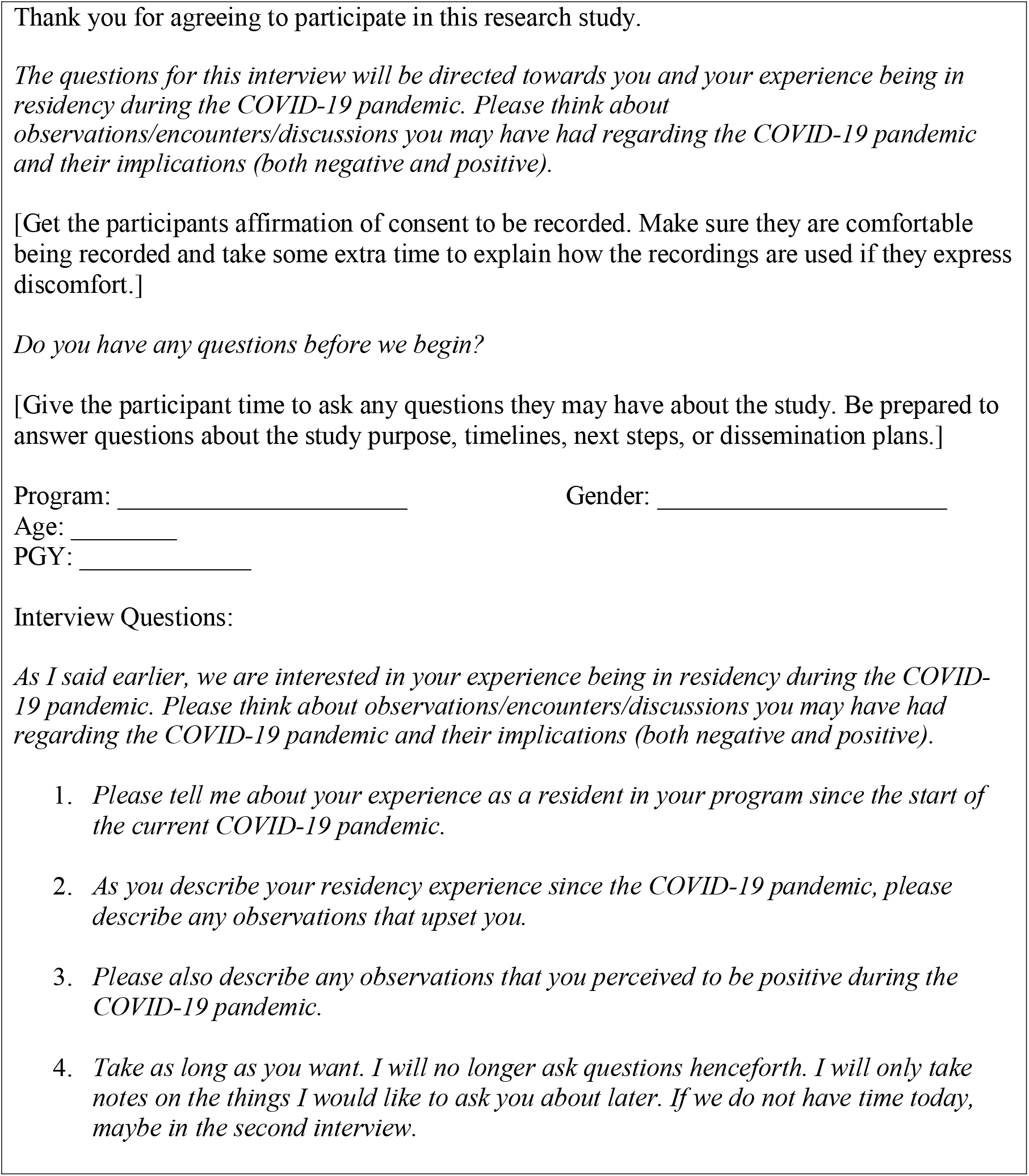
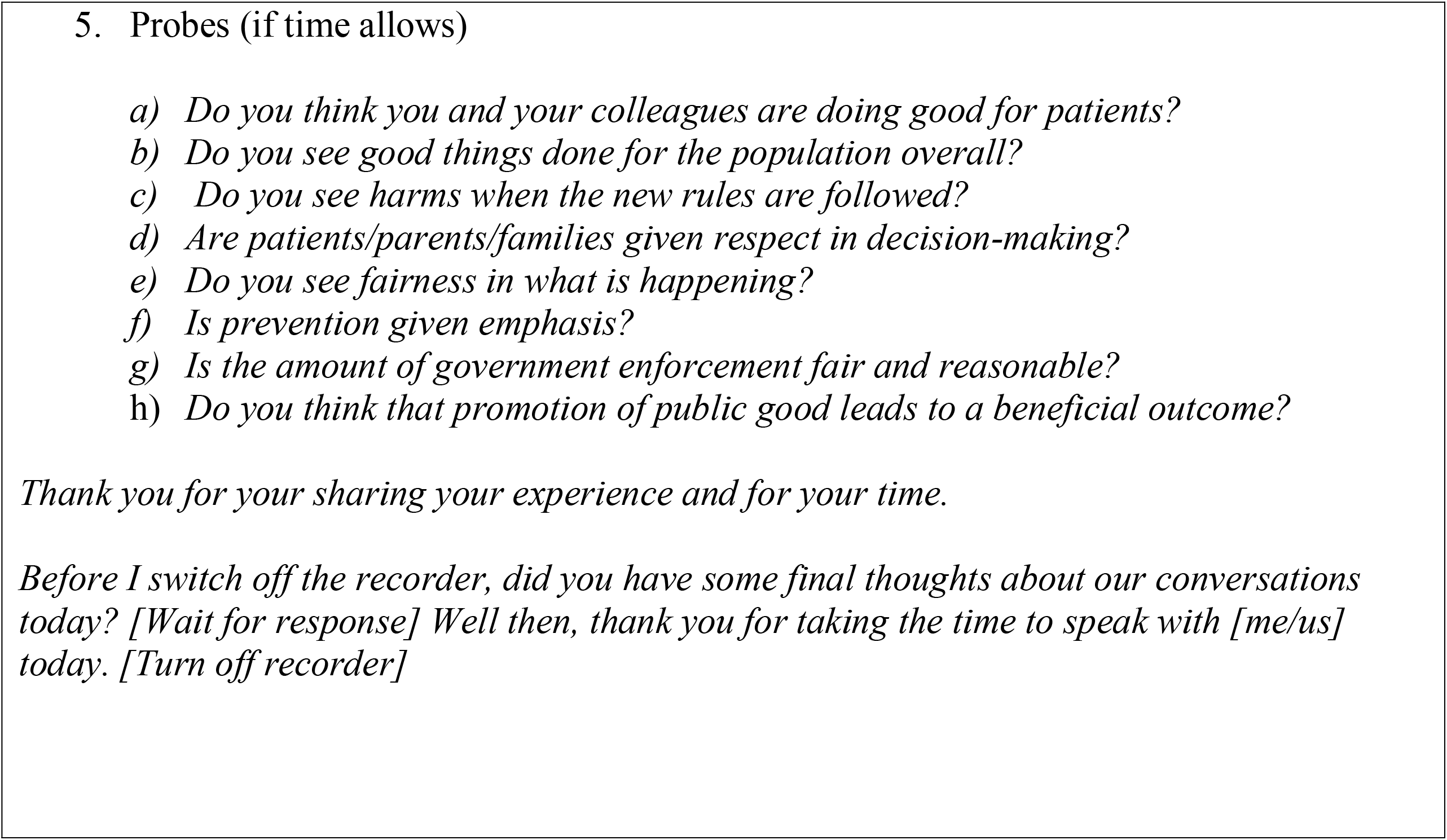
Interview Guide.

### Data Analysis

The interview data was analyzed in several ways. To construct the main themes in which ethical issues from the resident physician stories were determined, we used the six steps of thematic analysis as outlined by Braun and Clark’s framework. ^33 34^ These steps involve: becoming familiar with the data; generating initial codes forming sub-themes; searching for main themes; reviewing these themes; defining the main themes and writing the themes in a coherent document. Transcripts were de-identified and sent to the research team (three co-authors) for independent review. Emergent themes and their definitions were discussed with the co-authors until consensus was reached. Ethical issues suggested by the stories and by the researchers were described as occurring within each theme.

From a narrative inquiry perspective, the researcher first interviewed the participants, then analyzed the participant stories for threads. Threads differ from themes because they “attend to experience across lives. Rather than reduce experience into categories or themes, the concept of threads understands the complexity of experience in entirety…” ^26^

By acknowledging threads, the researcher wove together the stories which is “more responsive to the complexities of the lives of the participants.” ^26^ Furthermore, threads move across the themes found in the participant stories as described above.^24 25 28^

The process of narrative inquiry is mostly interpretive, steered by what the researcher sees as resulting from the data; the subjective role of the researcher is central to the process. ^24 25 28^ Furthermore, as part of NI, and to protect the anonymity of the participants, a composite story was constructed to represent a collective story of the milieu of residents’ stories and the themes characterized by the researchers. A composite story is a data analysis technique that “allows the weaving of the narrative resonant threads within each individual narrative account into a story that encompasses the voice of the participants.” ^35^ The composite story becomes the collective story creating the final research text which is the narrative analysis of the story. ^36^ Thus, in our inquiry, to protect the identity of the residents, we selected *Junior* (personal pronoun they/them) to represent the residents’ experiences and responses. The collective story is presented as a journal entry by *Junior* comprised of direct quotes which were edited for brevity and to exclude possible identifiers.

## Results

Telephone interviews were held with eleven resident physicians who were in their first year of training. Interviews lasted on average 45 minutes, and all the questions, including probe questions, (see Table 1 with interview guide) were asked. The age of participants ranged from 25 years to 30 years. Of the residents, n=5 identified as women and n=6 identified as men. Residents were recruited from Internal Medicine (n=2), Family Medicine (n=2), Ophthalmology (n=1), General Surgery (n=1), Pediatrics (n=1), Diagnostic Radiology (n=1), Public Health and Preventative Medicine (n=1), Psychiatry (n=1) and Emergency Medicine (n=1).

There were three main themes that were suggested by the participants’ stories of their experiences during the first wave of COVID-19. These main themes were: 1) *Intersecting healthcare and medical education systems*, 2) *Public health and the public good versus the individual*, 3) *Health systems planning/healthcare delivery*. For these themes, the stories suggested ethical issues within the context of the first wave of the COVID-19 pandemic as described below.

### Intersecting healthcare and medical education systems

Residents described having little to no autonomy and agency with regards to their own learning and working environments, which impacted their personal wellbeing, patient wellbeing (or both), and may have led to moral distress. Indeed, this theme of intersecting systems was not unique to the pandemic but may have been worsened by it. We perceived this to be an ethical issue pertaining to respect for autonomy and fairness.

> *I was being pulled off my home rotation which is something I wanted to be on. And then, being asked to do two more months of an intense medicine rotation back-to-back, you know, I – I didn’t feel super comfortable bringing up my concerns because, at the same time as a physician, as a resident I want to be there for patients or want to be there to do my duty in these times. So that was hard, especially being so junior just with the power differential with, attendings and so on. It was kind of that emotional and mental vulnerability. So, a lot of us, got re-deployed and it felt like our voices weren’t quite as heard. We were the most vulnerable and then we just kind of had to go wherever PGME wanted us to go. 011*

An ethical issue suggested by the stories was the paradox of the resident’s role as a novice learner while simultaneously becoming a major part of the healthcare workforce. While this potential ethical issue existed prior to the pandemic, the pandemic may have made this paradox more prominent. This ethical issue encompasses principles of beneficence and non-maleficence, balancing both patient and resident physician needs. While being re-deployed was voluntary, resident physicians reported harm to their wellbeing because of the increased workload.

> *So, the three residents that were on this particular rotation all got scheduled for extra calls. When I asked the Program Administrator about this, essentially all they did was carbon copy the Program Director in an email and put in the new PGME guidelines that said because of the pandemic the call was allowed to be scheduled in theory as an indefinite number of calls. It just ended up being one more thing for each of us and it essentially silenced anything a resident could do. Again, it’s not an issue with being happy to help, I just feel that a lot of decisions have been made on our behalf without us being present. And all those things just make it more difficult for you to continue to be well. 003*

Residents also reported a lack of communication and transparency in how decisions were made about their training and work. They also reported that they were not engaged as key stakeholders in the planning process for healthcare workforce planning and training disruptions. This too highlighted principles of beneficence and non-maleficence.

> *The healthcare system and PGME were making these changes very suddenly, on a reactive basis, and you are setting precedence for what could happen also in the future. I guess you don’t really have a lot of time to think about it … or we don’t really have a say in it, because … those rules are already being made. As a resident you don’t have a say at all. 010*

As part of this, residents often felt ill-prepared and uncertain about how to navigate the ever-changing circumstances of learning and working, and whether to give up learning opportunities for the sake of personal safety and vice-versa.

> *Could I protect my family if I had had any kind of potential exposure? The situation always became, well, depending on the staff, the residents could see the patients, but then honestly, you’re putting yourself at higher risk. Or you don’t see the patients and only those who need to go in the room, like staff or perhaps a senior resident in that service would go in, and you lose out on the learning opportunity, but then you have a slightly better chance of protecting your family or the people you live with. I guess that was always a tough situation that happened more than once. It made me feel like I had to pick and choose. It made me feel like it was an unfair decision. Because obviously I would want to protect my loved ones, and obviously I would want to care for my patients. So, it felt like an impossible situation. I guess I defaulted to the decision of what would be the best, safest thing to do for everyone involved and that would be giving up a learning experience and have limited people treating the patients in the room. Because the staff and/or senior would have had to see the patient, anyway since I was a junior off service resident. So, it made more sense to give up the learning experience, have a better chance of protecting my own loved ones and also getting the right care for the patient. I guess it was just sad that I had to make that choice. 009*

In contrast to the above issue of giving up a learning experience for personal safety, one resident felt underutilized and as though they were missing out on training opportunities despite their awareness of the safety risks. On the one hand, residents were being protected as learners, but on the other hand they were not able to engage in formative learning opportunities that were important within the context of CBME and for patient care.

> *I think the most upsetting part was maybe when it came to limiting residents’ participation in sick patients. I can only speak to my discipline, but in my discipline, we were no longer able to be part of the resuscitation of the very sick patients. That is obviously with the thought to protect residents. I think uniformly that was good policy. But I do think the difficult aspect that some of us had was not being able to participate in these patients’ care while it may be beneficial for us as something that is part of our training. Not being involved in those scenarios felt like it was a loss of our skill utilization or application of our skills and a learning opportunity. I think a lot of the resident roles and what our duties were basically from a top-down approach, where the guidelines were set by PGME. I don’t know if this happened behind the scenes, but to my knowledge there wasn’t a lot of resident participation in the sense of what we would prefer to do, what we feel comfortable doing, how to best utilize our resources. 008*

### Public health and the public good versus the individual

Residents reported a struggle between balancing individual patient needs with the needs of the broader public. One example was the visitation restrictions of non-COVID-19 patients in pediatric intensive care, as described by participant 7 below:

> *We had visitation regulations and rightly so. But with children, we really try and have both parents in the room, if children have both parents involved in their care because they can help support each other. But with COVID-19 our restrictions, only one parent was allowed in the room, and there wasn’t any switching around, and they weren’t allowed to leave the room if anyone was COVID pending, like, for any reason. So, it’s hard, because you can understand where those politics came from, because it helps limit the spread, but we had some cases where we had to inform families that their child was palliative, or it was an end-of life discussion, where it was a child with COVID-19 and you were talking about withdrawing care. Having only one parent involved in those discussions, just honestly felt inhumane. A lot of times we ended up just saying, you know what, both parents can be in the room, like, they both need to be with their kid. This is something that would impact the rest of their lives. But it’s hard, and it’s even hard from the standpoint of with personal protective equipment, it’s really hard to connect with patients, because you’re wearing a mask. So, a lot of these difficult and crucial conversations that we will have with families about goals of care and about end-of-life. When you’re talking about someone’s child, doing so behind a mask just feels very cold and distant. It’s just so hard. All of these things were done for very good reasons, but seeing the impacts that it had on some of these families, and I think particularly because we didn’t see as many COVID cases at the Children’s Hospital. Overall, the hospital wasn’t as affected, which of course may be in part because of these policies. It’s just so hard, and it’s hard to see that very real, very human impact, and that impact on those families and balancing having good quality care in the sense of having a humane coping strategy for children and families and having that support there, versus the public health strategy of limiting the spread of COVID-19. 007*

We identified an ethical issue concerning visiting regulations for palliative care and developing rapport with severely ill patients whilst wearing personal and protective equipment. On the one hand, personal safety for both the patient and resident is important, yet on the other hand developing rapport during a patient’s time of need, such as their end-of-life, is also just as important.

> *They’re in an unfamiliar environment, strange noises, and you know with everybody in the healthcare team having to wear a mask, gloves, you can’t see our faces, so everybody looks similar so it’s hard to see who’s coming to visit the patient. Patients struggled with that and being alone, which was upsetting, because some of those patients were admitted in a terminal capacity with complications from their diagnosis that would end up killing them within a short period of time. I think one of the saddest experiences I had was, we had a dying patient on a particular ward and the patient’s spouse was sitting at the bedside holding their hand and I was on call, I was just making the rounds to make sure that the patient was still comfortable, that the team was there in case anything else needed to be done, if I could offer perhaps anything else, and the spouse said that the patient was comfortable, didn’t have anything else off the top of their head that they needed, but it was really the look on their face that was tough to see because you could tell that they were totally lost. In that person’s most desperate hour when they were trying to be the support person for their dying spouse, they themselves had nobody else to support them in that moment. That was certainly tough. 006*

Residents also reported a lack of guidance from education and healthcare leadership with regards to patient safety issues that could arise over virtual appointments with patients. For example, they reported having to diagnose patient illnesses over the phone, as opposed to in person in order to protect their own personal safety and that of the public. This virtual form of diagnosis superseded the need to protect and treat the individual patient.

> *I think we’re trying to do the best for our patients, but we can’t see many of them in person because they might be at high risk, or they might be too scared to come in. I think we’re still able to help many of our patients through giving them general advice about COVID-19, through general medicine over the phone and still trying to meet many of their medical needs over the phone, or over a video-call. I think there are a lot of instances where we’re too limited by what we can do right now through virtual care. I’m sure there’s probably a lot of things that fall through the cracks because of these different limitations that we’re not used to. For example, having to diagnose patients’ rashes over the phone, or even through pictures that they try to send us. I’m sure there’s probably lots of instances where we probably could have done a lot better job if we were to see the patient, and as a result I was possibly not treating them properly because we were not able to fully diagnose them in person. 001*

### Health systems planning/healthcare delivery

Another theme involved health system planning and healthcare delivery by medical learners, a group which includes both residents and medical students (clerks). In certain specialties, the removal of clerks and off-service residents from clinical units treating COVID-19 patients led to a disproportionate load on the remaining residents from particular specialties such as internal medicine. This disparity may have led to unfairness and was apparent in areas such as cross coverage of being on call, for example. However, clerks were removed to protect their own safety.

> *I guess the pandemic started and the clerks were being pulled back. I can only speak from the level that I’m at. It became obvious to us how dependent the medical system was on medical students for coverage, especially on the internal medicine ward and for those of us who were on different subspecialties. So essentially with the cross coverage, not only does it become more difficult for teams that you don’t know about, but also when a consult comes in essentially, it’s being broken down and split across three people and you only have two people, so effectively your night is difficult and you get no sleep at all. I think the hardest situation for patients is that I don’t know anything. There was a senior obviously, and nothing against them, but it’s just an issue of the lack of labour and the fact that there are so many mental processes going on that it’s easy to forget and you’re obviously triaging so many things as you go. I just find myself frustrated with this whole systemic issue about how we never feel that we’ll ever be adequately serviced. COVID-19 just made that much more apparent. 003*

Residents reported the experience of having resources allocated towards COVID-19 surge preparation, while services to patients with other healthcare needs were restricted. We saw this as a potential ethical issue related to fairness because resources may have been unfairly allocated, yet it was important to prepare for a possible surge in COVID-19 cases to prevent further harm to the public.

> *I think that probably the greatest thing that I found difficult, was trying to reconcile the nature of walking through our hospitals whilst they were largely emptied in anticipation of a real surge, and then hearing the stories of patients presenting late and having advanced pathologies, because they were too concerned to come into hospital. 005*

Another example of an ethical issue suggested by the data was the allocation of resources to areas of healthcare outside of public health mechanisms. Again, this might be considered an ethical issue around fairness.

> *In terms of emergency preparedness and emergency response we noticed that the clinical acute care side of medicine mobilized relatively quickly compared to other areas of the system. Medical students were asked to volunteer as contact tracers so there seemed to be a disparity of resources. 002*

A further issue related to health systems planning and healthcare delivery was that many residents were working long hours without breaks. These residents received little reprieve from staff physicians as well as minimal acknowledgement of both their work contributions and the self-sacrifices they made with respect to their own education/training opportunities. While the issue described below may not necessarily be wrong because electives were cancelled for both learner and patient safety reasons, this resident recognized the need to make pragmatic decisions during a time of uncertainty.

> *I mean is it individually fair that a lot of people, had electives taken away from them? (They had opportunities to visit other centres where their career may take them*.*) No, I don’t think that’s fair, but was it needed? Yes, I think it was. But not being able to go on a much-needed vacation, and the burnout that there is for staff members and for healthcare workers to be put in situations that were potentially dangerous for themselves…. I think a lot of us sacrificed a lot and granted, some of it was not necessarily a voluntary sacrifice … but I don’t think that there was any choice in the matter really, with people having to give up going to those kinds of places for electives. Even the clerks for instance having to put off a lot of the rotations, it’s not the choice that they necessarily would’ve made. But I don’t think it was unfair; I just think it was unfortunate. 004*
>
> Another example of an ethical issue with respect to fairness and respect is that residents reported feeling a lack of appreciation from the healthcare system, as they only received a “thank you letter” since re-deployment was voluntary. Again, residents felt a lack of acknowledgement in health systems planning and healthcare delivery despite being integral to both.
>
> *Essentially all we got is a thank you letter at the end of the day. Again, not that I’m saying that there should be a reward, but even to take a breath, all of these things are taking away from being able to. All of our social events have been taken away for a good reason. I feel that the system didn’t do anything to protect us, all it did is spread us thinner. I’m not familiar with any other staff coming onboard. The only things that happened were staff covering the COVID teams, but the COVID teams were not the issue because my understanding is that they were not that busy. I don’t feel that we got that much help at the end of the day. 003*

Amidst the ethical issues we identified above regarding autonomy and being engaged to prevent harm to patients and the residents themselves, there were positive aspects to the disruption caused by the COVID-19 pandemic that arose from the participant stories. These encompassed the notion of altruism and understanding the duty to treat despite re-deployment being voluntary and uncompensated. Participant stories also noted that COVID-19 brought healthcare providers together, highlighting the importance of teamwork, collaboration, and solidarity (both within healthcare and within the broader community). Three key narrative threads resulted from resident stories overall, which highlighted their journey during the first wave of the COVID-19 pandemic. They were: 1) *Engage us, 2) Because we see the need for the duty to treat, 3) And we are all in this together*. These threads move across the themes found in the participant stories. Table 2 shows an example of how the threads move across the theme of wellbeing of the learner, patient or both learner and patient.

**Table 2.**
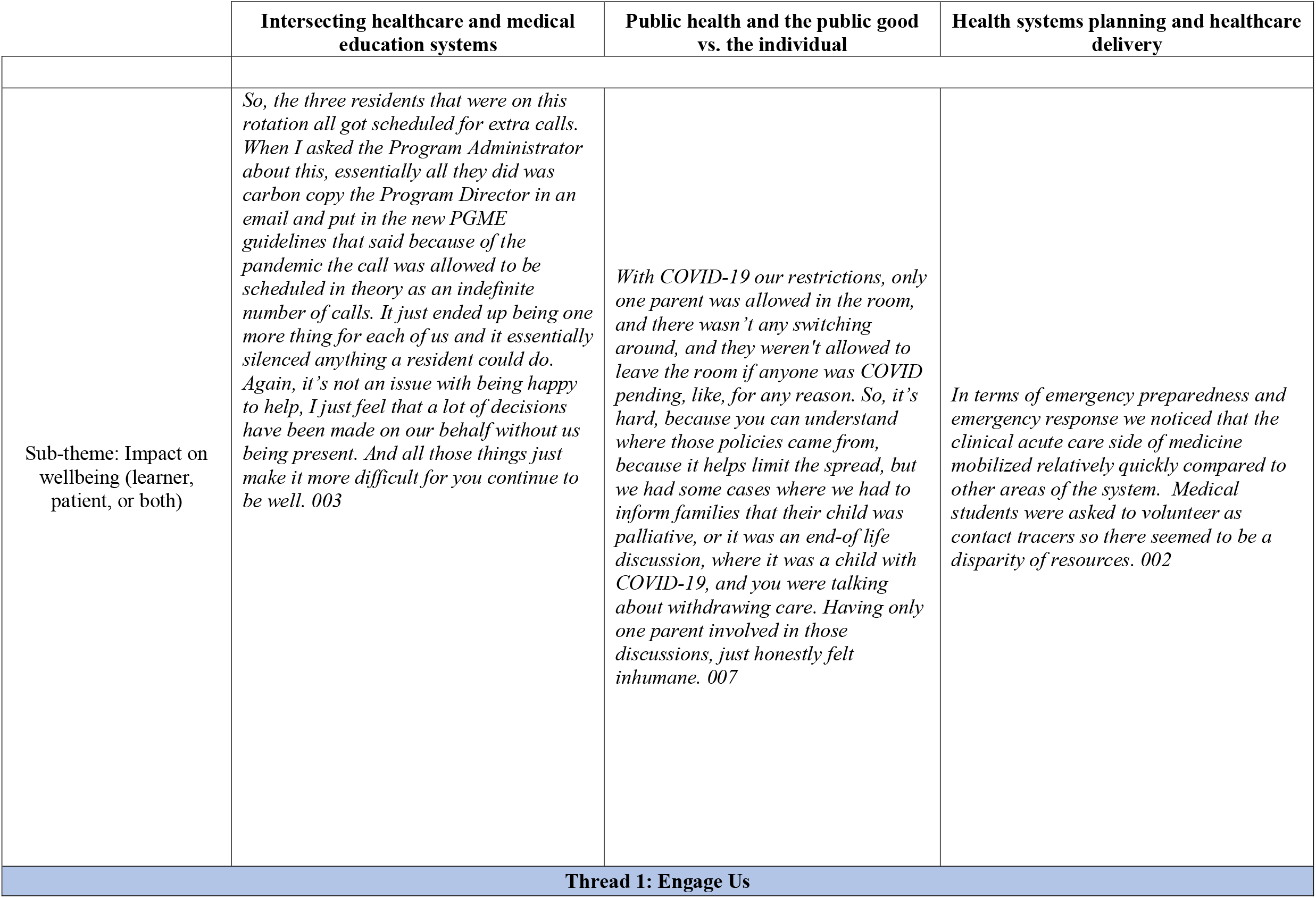
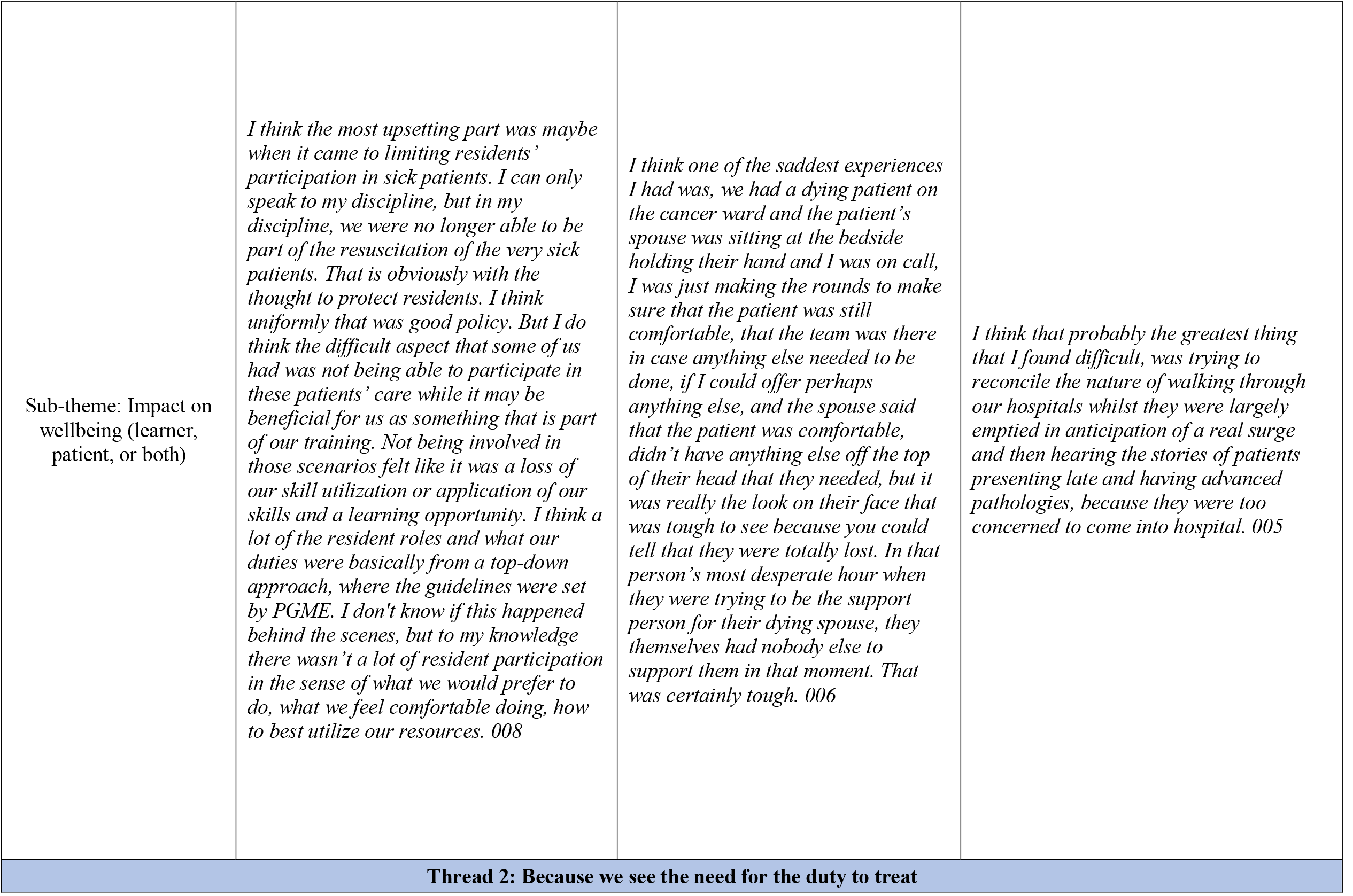
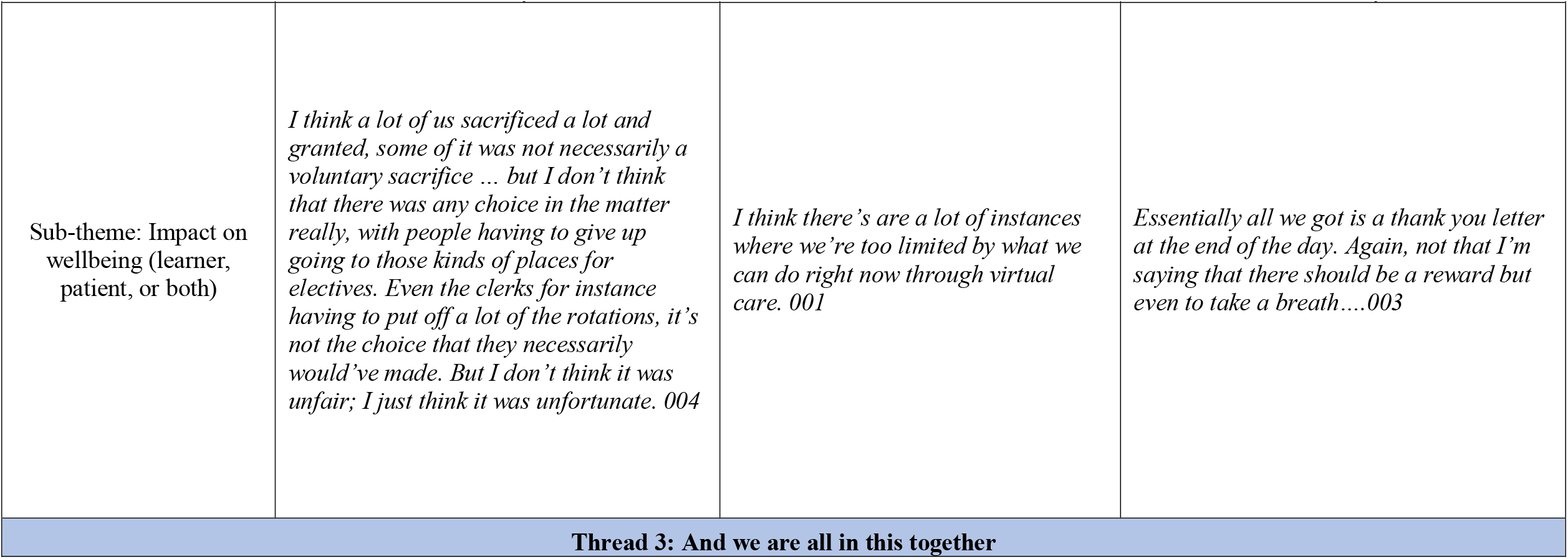
Example of threads present across sub-theme and main themes.

A composite story within which the threads tie both the negative (challenges) in which ethical issues arose together with the positive aspects of the pandemic. Table 3 shows this as a composite of all eleven residents’ stories.

**Table 3.**
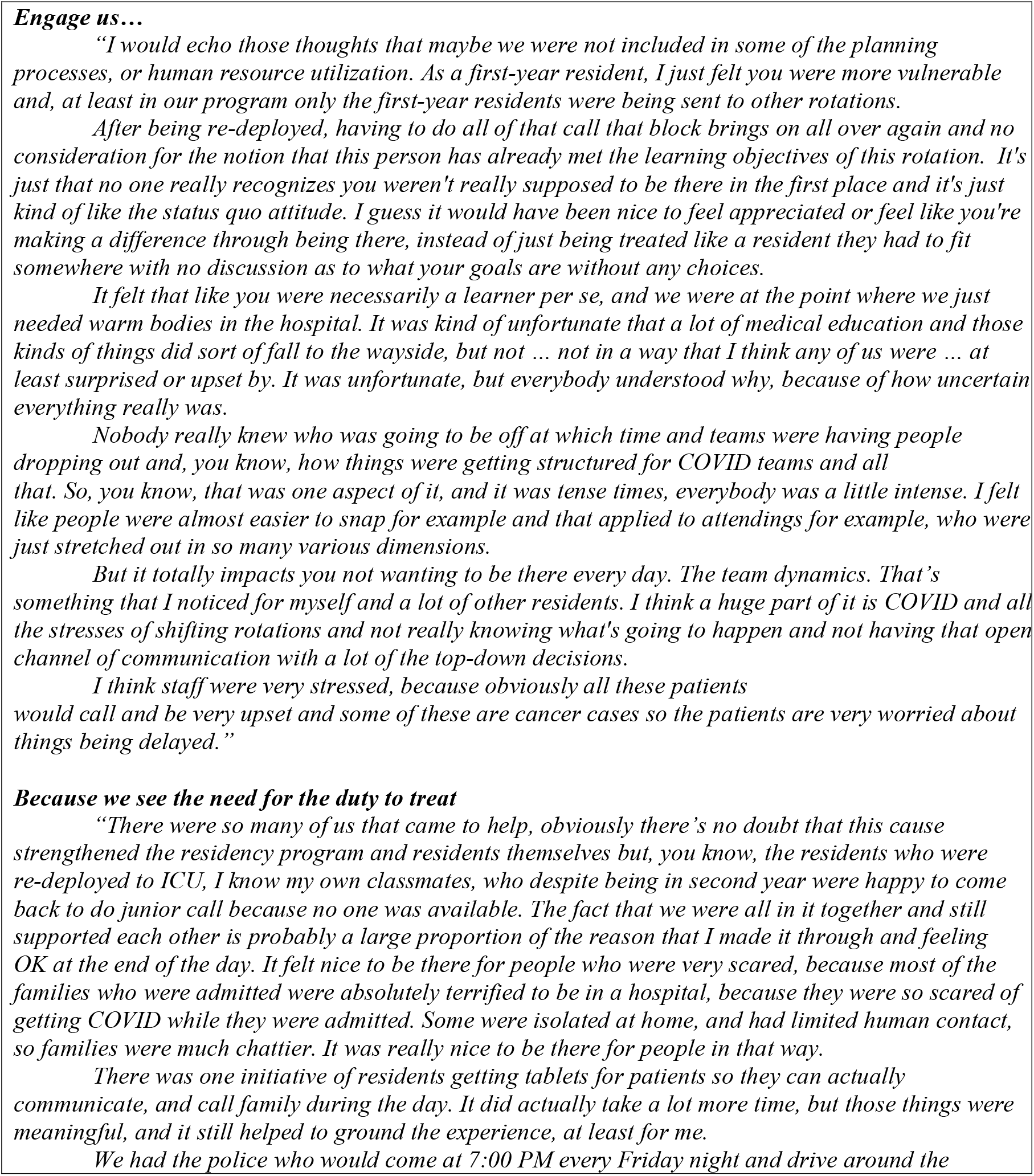
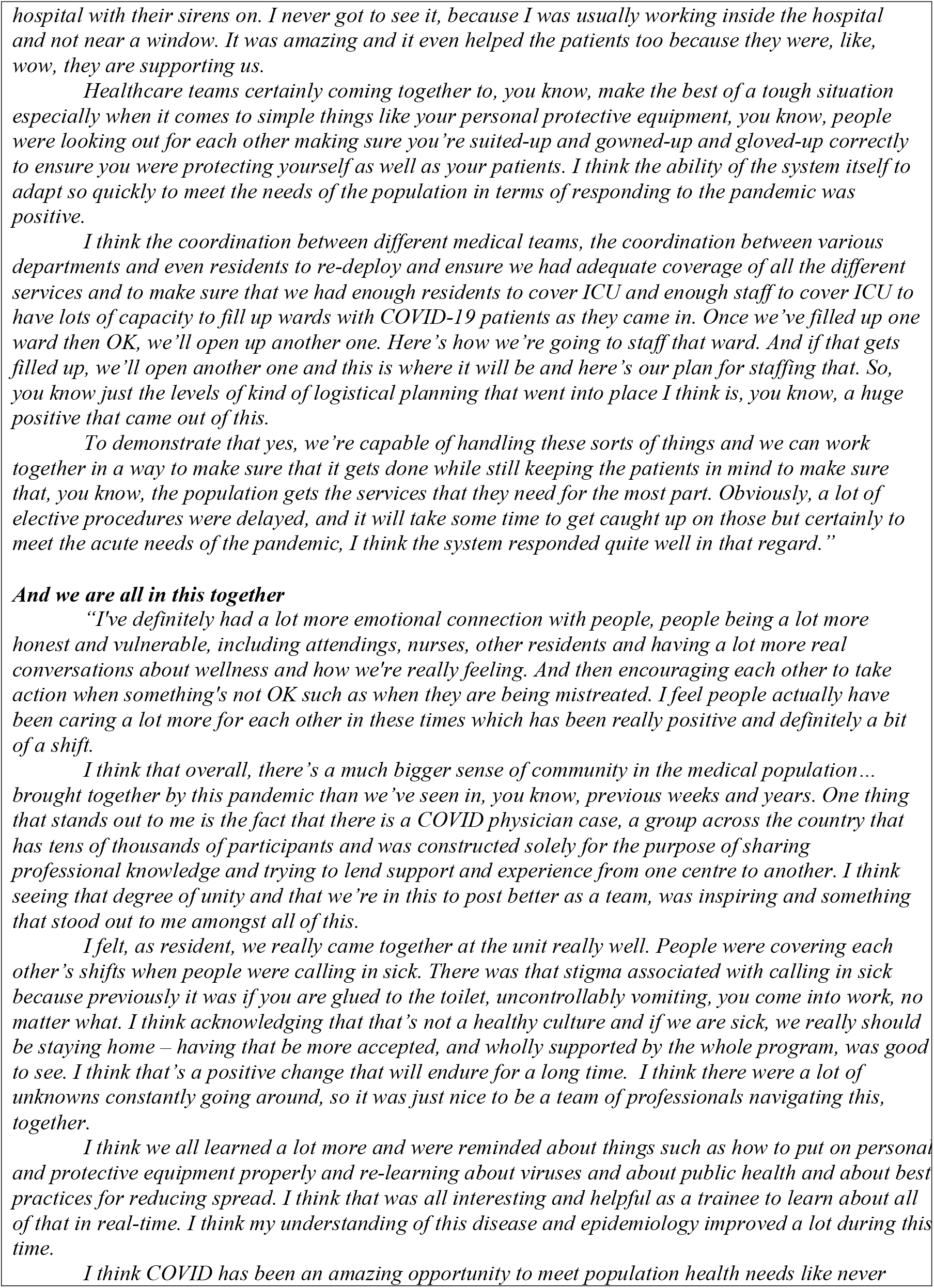
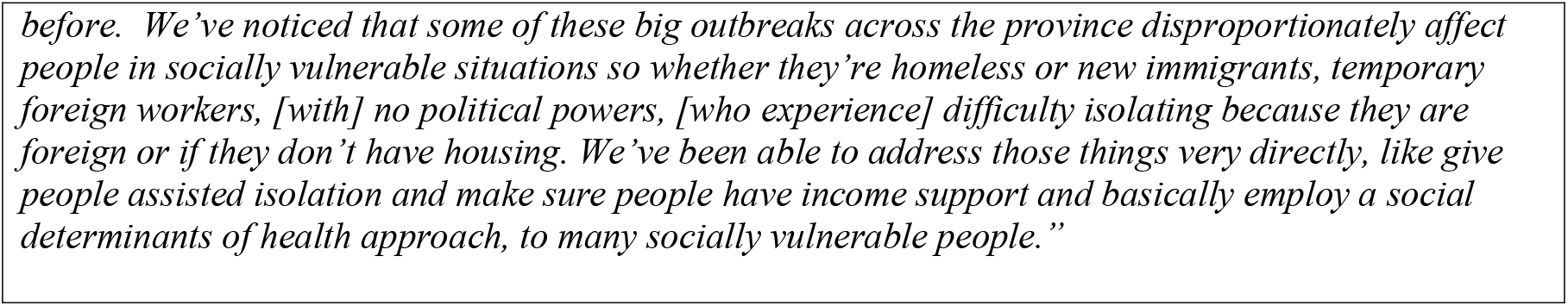
Junior’s Journal Entry.

## Discussion

These narratives provide a “window” ^37^ into junior residents’ experiences of the first wave of the COVID-19 pandemic. We captured a unique set of cross-sectional data from first year residents enrolled in various residency training programs who were willing to share their experiences during a time of stress, moral distress, and uncertainty. Their stories were emotional and reflective. Through these narratives, we constructed potential ethical issues that may have arisen. This study illustrates the complexities of residency education during the COVID-19 pandemic. The junior residents explained their experiences, emotions and particularly outlined changes in their learning and working situations. ^38^ Their reflections on their situation, that of being both involved or uninvolved in direct patient care, were numerous and sometimes conflicting. Nevertheless, they were able to articulate not only the challenges, but also the opportunities that each situation presented in a way that allowed them to see the bigger picture of a macroethical situation that is, protecting the public good. The residents expressed this unease between frustration and achievement, disappointment, and gratitude amidst sudden changes in their work and training situations that they had little or no control over.

Moreover, the impact of the COVID-19 pandemic has subjected all healthcare providers to previously unchartered territory where they were faced with difficult circumstances and potentially life-threatening work conditions. In particular, they were forced to balance their own wellbeing with that of their patients, while providing care for all unwell patients within a healthcare system that had insufficient resources. ^39^

### Engage us

The literature has shown gaps existed in ethics education prior to the pandemic ^40 41^ and this deficit may have likely been exacerbated by the pandemic. Prior to the pandemic, residents were working in microethical situations surrounding commitment to the profession, commitment to patients and commitment to their own wellbeing as per the Professional Role, but with little connection to commitment to the public.

During the pandemic however, commitment to the public good (a macroethical situation) encompassed all microethical dealings of being a learner and a healthcare provider which in turn impacted commitment to the profession, to patients as well as to themselves and their wellbeing. Residents thus have had to navigate both microethical and macroethical situations whilst being both a learner and healthcare provider, a scenario which may have resulted in moral distress.

### Because we see the need for the duty to treat

Our findings of a perceived lack of autonomy may lead to a perceived lack of agency and powerlessness, despite the voluntary nature of re-deployment of residents to COVID-19 pandemic related work. Although residents showed altruism, there was a tension from their stories that indicated that altruism could diminish their wellbeing as a result of an increased workload and a lack of appreciation. Zaidi et al., advocate for the application of relational (care) ethics ^42 43^ within the context of the COVID-19 pandemic by healthcare leadership that would protect healthcare providers such as residents from “self-sacrifice.” As such “non-maleficence could be expanded to allow for certain types of interventions, i.e., not just refraining from doing harm but actively interfering or taking action if wrong is being witnessed” between patients and healthcare providers, or between healthcare providers alike. They go on to state: “if we are truly concerned about the wellbeing of healthcare providers, the principle of beneficence should be restricted to prevent the systematic self-sacrifice that results in harm.” ^44^ This means that the culture of healthcare more broadly must balance the notion of harm against the patient and harm against the healthcare provider, especially during times of uncertainty such as the COVID-19 pandemic. This itself is an ethical issue because how, where, or to whom should the principle of beneficence be restricted? And, at what point should healthcare providers not put patients above themselves? This issue becomes even more complex when the healthcare providers are resident physicians who comprise a large part of the healthcare workforce but are still considered learners. Moreover, the context of a global pandemic further complicates this issue. ^45^

### And we are all in this together

With respect to public health, the relationship between the principles of clinical ethics ^46^ such as autonomy, beneficence, maleficence, and justice may warrant further examination in residency education within the context of the COVID-19 pandemic and future public health emergencies. Residents identified the positive aspects of teamwork, collaboration and solidarity that grew from the pandemic and were evident to have developed between healthcare providers themselves, as well as between healthcare providers and the broader public. The notion of protecting the public good and the prevention of harm became the forefront of healthcare during the COVID-19 pandemic. However, to what extent does protecting the public good allow collective beneficence? Indeed, what ought to matter is a shared interest in survival, safety, and security—an interest that can be effectively pursued through the pursuit of public goods and through ongoing efforts to identify. At the same time though, consideration of privilege and disadvantage in particular individuals or groups within the greater population could still foster an ‘us’ versus ‘them’ divide when bearing in mind the public good. ^43^

This highlights the importance of working together towards a common goal of wellbeing ^19^, yet recognizing that wellbeing may be conceptualized differently by individuals and social groups/programs. This is itself an ethical issue and would be important for consideration at the systems level of PGME as well as health systems planning/healthcare delivery. ^47 48^

For example, is there a dominant group that is at a particular disadvantage when considering the public good? Does a collectivist approach undermine and marginalize individuals or social groups to the point of non-maleficence? To help mitigate the collectivist approach taken by our institution, the PGME office is convening a COVID-19 debrief committee which will engage residents to determine areas for improvement in pandemic planning for the future.

### Future directions

The key differences between public health ethics and clinical ethics can be understood by the unique nature of public health and summarized by four main constructs inherent to public health ethics: (a) public or collective good; (b) the promotion of public good with specific emphasis on prevention; (c) the promotion of public good and the role of government enforcement; and (d) the implicit notion that promotion of public good leads to a beneficial outcome. ^19^ While the four principles can be applied to the public health constructs above, the backdrop of a microethical issue changes to that of macroethical issue amidst a pandemic. Consequently, the entity of concern shifts from the individual to the greater public good which may require education around how the four principles may change and how ethical issues arising between the individual versus the public might be navigated.

More specifically we acknowledge the importance of considering both microethics and macroethics ^13 15 18^ at the systemic levels of public health, as well as within vulnerable populations such as resident physicians, patients, and their intersections. Junior resident physicians and patients may have compromised autonomy because they may lack in agency due to their lower status in social power and the hierarchy of medicine. Alternatively, and given they are learners and unwell respectively, they could be considered vulnerable since their safety and protection was also a priority for leaders in residency education and healthcare alike. ^49^ Thus, leadership may not expect junior residents to be engaged in the decision-making process ^50 51^ during a public health crisis such as the COVID-19 pandemic as they are part of the larger population to be protected from harm. This was shown through the potential ethical issues we found as well as the resident stories overall.

The ethical issues arising from the first wave of the COVID-19 pandemic and constructed from the stories told by resident physicians may serve as a foundation on which ethics teaching and future pandemic planning can take place. Gaps in ethics teaching existed prior to the pandemic, and the pandemic has subsequently highlighted the need to examine how the clinical ethics principles may shift during public health emergencies.

Future teaching and research may benefit from considering both macroethics and microethics (and how and where they intersect) in curriculum development. This approach would ensure residents have the conceptual tools to deal with both patient-physician interactions (microethics) and physician-public health interactions (macroethics). Furthermore, as part of residency education in a broader sense, an emphasis on declaring and engaging all key stakeholders during pandemic and disaster planning would also better equip PGME and the healthcare systems.

There are several limitations to this study. First, it does not take into consideration the political climate of the region at the time of data collection. From a public health perspective, the socio-political structures influencing systemic decision-making during the pandemic warrants further examination when considering the ethical issues reported in this study. Second, this study was conducted during the first wave of the COVID-19 pandemic during which decisions were being made pragmatically and quickly due to many unknowns about the virus, hence these ethical issues may not be relevant to subsequent waves of the pandemic.

## Conclusion

Our study is the first to attempt to understand junior resident experiences during the first wave of the COVID-19 pandemic. Despite challenges, there were positive aspects to the pandemic such as teamwork and collaboration as well as the notion of altruism carried by residents, many of whom volunteered to help achieve what was best for the public good. Efforts to understand how resident physicians can be engaged in their own education as well as how to navigate public health crises with respect to ethical principles in both microethical and macroethical situations could benefit both residency education and healthcare delivery alike.

## Data Availability

The participants of this study did not give written consent for their data to be shared publicly, so due to the sensitive nature of the research supporting data is not available.

## Footnotes

1 Populations are constituted by diverse communities of heterogeneous beliefs and practices. They represent communities and the broader social and environmental influences of health on these communities collectively. In this manuscript, populations refers to the whole number of people in each region.

